# Cumulative incidence of SARS-CoV-2 and associated risk factors among healthcare workers in the Eastern Cape, South Africa

**DOI:** 10.1101/2021.11.08.21265966

**Authors:** David Stead, Oladele Vincent Adeniyi, Mandisa Singata-Madliki, Shareef Abrahams, Joanne Batting, Eloise Jelliman, Andrew Parrish

## Abstract

**Objectives:** This study assesses the cumulative incidence of SARS-CoV-2 infection among healthcare workers (HCWs) during South Africa’s first wave and examines the associated demographic, health-related, and occupational risk factors for infection.

**Methods:** Multi-stage cluster sampling was used in a cross-sectional study to recruit 1,309 HCWs from two academic hospitals in the Eastern Cape, South Africa over six weeks in November and December 2020. Prior test results for SARS-CoV-2 polymerase chain reaction (PCR) and participants’ characteristics were recorded while a blood sample was drawn for detection of IgG antibodies against SARS-CoV-2 nucleocapsid protein. The primary outcome measure was the SARS-CoV-2 cumulative incidence rate, defined as the combined total of positive results for either PCR or IgG antibodies, divided by the total sample. The secondary outcome was significant risk factors associated with infection.

**Results:** Of the total participants included in the analysis (N=1295), the majority were female (81.5%), of black race (78.7%) and nurses (44.8%). A total of 390 (30.1%) HCWs had a positive SARS-CoV-2 PCR result and SARS-CoV-2 antibodies were detected in 488 (37.7%), yielding a cumulative incidence of 47.2% (n = 611). In the adjusted logistic regression model, being overweight (Adjusted odds ratio (AOR) = 2.15, 95% CI 1.44-3.20), obese (AOR = 1.37, 95% CI 1.02-1.85) and living with HIV (AOR = 1.78, 95% CI 1.38-were independently associated with SARS-CoV-2 infection. There was no significant difference in infection rates between high, medium and low COVID-19 exposure working environments.

**Conclusions:** The high SARS-CoV-2 cumulative incidence in the cohort was surprising this early in the epidemic and probably related to exposure both in and outside the hospitals. To mitigate the impact of SARS-CoV-2 among HCWs, infection prevention and control (IPC) strategies should target community transmission in addition to screening for HIV and metabolic conditions.

**Strengths and limitations of this study:**

- This is a large representative sample of the total workforce of the two hospitals, with a good spectrum of staff category.
- Combining the historical SARS-CoV-2 PCR results with the Nucleocapsid IgG enabled capturing of some of the asymptomatic and missed SARS-CoV-2 infections.
- This is one of the first studies to look at SARS-CoV-2 infection risk factors in a high exposure environment in Africa.
- A limitation is that HIV ELISA and CD4 counts were not tested, but relied on self-report, which may likely underestimate the burden of HIV in the cohort.

## Background

South Africa reported its first imported case of SARS-CoV-2 on 5 March 2020 and subsequently experienced high rates of transmission throughout the country. The first wave peaked in July 2020, the second wave in late December 2020 and a third wave occurred in June 2021, with total cases approaching 3 million.^1^ The Eastern Cape ranked 4th out of South Africa’s nine provinces for cumulative SARS-CoV-2 cases, with 290 898 cases recorded on 2 October 2021.^1^

Healthcare workers (HCWs) are responsible for providing acute in-hospital care for patients with moderate and severe COVID-19 who require oxygen support and other therapies.^2^ The HCWs are exposed to infectious droplets and aerosols, putting them at increased risk for infection.^2^ Despite infection prevention and control measures at the health facility level, HCWs still acquire SARS-CoV-2 at a higher rate than the general population.^2–4^ A prospective study of 200 frontline HCWs in the United Kingdom (UK), during the first peak of viral transmission involving the collection of twice weekly nasopharyngeal swabs for reverse transcription polymerase chain reaction (RT-PCR) and monthly blood samples for serology, showed that 44% became infected. This was more than double the rate of the local population.^3^ A smartphone application allowing self-reporting of positive SARS-CoV-2 PCR results was used in a survey of almost 100,000 UK and United States (US) HCWs. Incident cases in these HCWs were almost 12-fold greater than in a two million comparator sample of the general population.^2^ Another UK study found a SARS-CoV-2 seroprevalence of 16.3% among HCWs compared to a 5.9% national community rate.^4^

Reported information on SARS-CoV-2 infections among HCWs in Africa is scanty. Two hundred and twenty-two HCWs from single South African paediatric unit were included in a global comparative seroprevalence study (recruited June to August 2020), with a seropositivity of 10.36% (95% CI: 7-15.07).^5^ A pre-print of a serosurvey of 500 HCWs in Blantyre, Malawi, reported a 12.3% positivity rate.^6^ The Eastern Cape Department of Health reported a total of 11,262 HCWs infected with SARS-CoV-2 by 18 February 2021, with 262 deaths (2.3% fatality rate). The highest infection rates were among state-employed doctors and nurses (18.2% and 22.3%, respectively) compared to a 2.8% for the province as a whole.^7^

The potentially high SARS-CoV-2 exposure environment in hospitals enables acquisition of data on infection rates and associated risk factors amongst HCWs, that can assist in understanding the dynamics of SARS-CoV-2 transmission and the efficacy of infection prevention and control measures. In some studies, high-exposure clinical areas such as Accident & Emergency Units, acute medical wards and intensive care units have been associated with increased HCW infections when compared to administrative or support service areas.^8–10^ Others have shown no difference between staff roles, suggesting that most infections were acquired outside of areas of patient contact, or outside of the hospital.^11,12^ Inadequate availability or faulty use of personal protective equipment (PPE) are both factors shown to increase the risk of infection.^2,13^ Male HCWs and those with at least one comorbidity also appear to have an increased risk of acquiring SARS-CoV-2 infection.^8,14^ Outside the healthcare environment, a study of 3,802 SARS-CoV-2 tests performed in the UK found that infection risk was increased by male gender, age 40-64 years, black ethnicity, lower socio-economic status, chronic kidney disease, and obesity. In this study, smokers had a lower risk of infection.^15^

SARS-CoV-2 is a global pandemic, but has affected individual countries and their health systems to varying degrees. Explanations for this include a complex interaction of population and genetic vulnerabilities, social mitigation behaviour, and health system interventions. Due to the paucity of evidence around the impact of SARS-CoV-2 on HCWs in Africa, this study was undertaken to gain insights in this setting. Frere and Cecilia Makiwane hospitals are both in the Eastern Cape Province in South Africa. This is an under-resourced province with a less robust healthcare system than that in some other provinces. Both facilities experienced high numbers of staff infections and absenteeism during the first wave of SARS-CoV-2, with considerable disruption to health service delivery. This study was conducted to assess the cumulative incidence of staff SARS-CoV-2 infections (symptomatic and asymptomatic), and their associated demographic, health-related, and occupational risk factors. Findings from the study may inform planning and improve IPC measures related to infections with SARS-CoV-2 and other respiratory viruses in the province.

## Methods

### Study design and settings

This observational cross-sectional study was conducted in two academic hospitals: Frere and Cecilia Makiwane, in the central region of the Eastern Cape, South Africa. Cecilia Makiwane is a regional hospital that provides levels one and two healthcare services to the residents of Buffalo City and the Amathole district. Frere hospital is a tertiary institution which serves as a referral hospital for four district municipalities: Buffalo City, Amathole, Chris Hani and Joe Gqabi. Together they serve a population of almost three million residents and have over 4,000 HCWs: doctors, nurses, pharmacists, allied workers and support staff (administration, laundry, kitchen and mortuary).^16^

### Re-organisation of hospitals during the ‘first wave’

At the onset of the first wave, local protocols were developed in accordance with the National Institute of Communicable Diseases Guidelines for the management of confirmed or suspected cases of COVID-19.^17^ Designated COVID-19 units were created from the existing emergency units of the two hospitals. All individuals meeting the criteria for ‘patient under investigation’ and/or confirmed cases of COVID-19 were directed to the designated area within the emergency unit, where triaging and clinical evaluations were performed by the attending clinicians. In both hospitals, patients meeting the criteria for admission based on the severity of their condition and/or co-morbidities were admitted into designated COVID-19 wards. Patients who presented in critical condition were admitted into the hospitals’ intensive care units. The head of the internal medicine department supervised in-patient admissions in both hospitals, thereby ensuring that the COVID-19 protocol was the same at both facilities. All clinicians working in the designated COVID-19 wards and emergency units received training on the effective use of PPE. In addition, the hospitals formed logistics committees comprising senior managers of the hospital to ensure a constant supply of PPE for use by all personnel caring for patients with COVID-19. The Occupational Health and Safety (OHS) unit of each hospital created a database of COVID-19 infection among its HCWs. HCWs were required to submit confirmation of a SARS-CoV-2 PCR positive result as evidence of diagnosis and permission to proceed with the mandatory isolation of 10–14 days, in accordance with the guidelines.^17^

### Participants

All categories of HCWs in the two hospitals were eligible to participate in the study. To ensure inclusivity of all HCWs, the study adopted a multi-stage cluster sampling technique. Risk profiles were categorised according to the exposure areas identified by Iversen et al.: ‘high risk’ if the HCWs worked in Accident & Emergency units, designated COVID-19 wards, and intensive care units (ICUs); ‘intermediate risk’ if HCWs worked in non-respiratory admission wards, outpatient departments (OPDs), and other clinical areas; and ‘low risk’ if the HCWs performed administrative tasks and other non-clinical duties.^8^ Prior to recruitment, mass sensitisation about the study was conducted through union leaders, departmental heads and clinical managers. In addition, a communique was circulated across the two hospitals to create awareness of the study. Each working area was allocated specific days to allow those on night shifts as well as those who were off-duty to participate with minimal interruption to service delivery. In addition, a central recruitment area was created in each of the two hospitals to cater for HCWs who might have missed the dates allocated by their departments. There was no sample size calculation performed, but rather as many staff recruited as possible within the budgeted time frame for the study. The study was implemented between 4 November and 18 December 2020. SARS-CoV-2 vaccination for HCWs in South Africa only became available in March 2021.

### Procedure

Each department/work area provided a dedicated station where HCWs completed a manual questionnaire and blood samples were drawn. Two research nurses and four assistants underwent training on the research process and study instrument over a three-day period prior to commencement of the study. The research nurses measured HCWs’ height and weight according to standard protocols. Venous blood samples (about 5 mL) were drawn by the trained research nurses using an aseptic technique. All blood samples were tested for the IgG antibodies against SARS-CoV-2 nucleocapsid protein by the National Health Laboratory Services in accordance with standard protocols.

To link the results of SARS-CoV-2 PCR tests recorded on the OHS databases with the SARS-CoV-2 IgG antibody tests, while maintaining confidentiality, a unique identifying number was used to encode the participants’ details (names, date of birth and area of work) in the research register, which was accessible only to the investigators. The questionnaire data for the study were captured on the REDCap^®^ online database of the South African Medical Research Council server.

### Main outcome measures

Serum samples were analysed on an Abbott ARCHITECT *i*1000SR instrument using the Abbott SARS-CoV-2 IgG assay in accordance with the manufacturer’s instructions. This is a chemiluminescent microparticle immunoassay (CMIA) for the qualitative detection of IgG against the SARS-CoV-2 nucleoprotein. Strength of response in relative light units reflects quantity of IgG present, and is compared to a calibrator to determine the calculated index (specimen/calibrator [S/C]) for a sample (with positive at 1.4 or greater).This assay has a specificity of 99.9% from 1020 pre-COVID-19 serum specimens and a sensitivity of 100% at 17 days after symptom onset and 13 days after PCR positivity.^18^

Seropositivity was categorised as a binary outcome: a positive result of SARS-CoV-2 IgG was considered as evidence of prior infection (humoral immune response), while a negative result was considered as either non-exposure or as a decayed (lost) immune response. Cumulative incidence: This was a combination of a SARS-CoV-2 diagnosis (positive SARS-CoV-2 PCR and/or positive SARS-CoV-2 IgG).

Missed SARS-CoV-2 infection: This was defined as seropositive SARS-CoV-2 IgG without any documented diagnosis of SARS-CoV-2. The latter included symptomatic individuals with negative SARS-CoV-2 PCR or who never tested and asymptomatic individuals who had not undergone PCR testing.

### Covariates

Sociodemographic and clinical covariates were included in this study. Age, sex, race, highest level of education, profession and smoking status, among others, were self-reported in the questionnaire. Age was categorised by decades for the multivariate analysis. Exposure risks (such as direct contact with patients with COVID-19) and training on the use of PPE were also obtained. Certain comorbidities (diabetes, hypertension, HIV, Tuberculosis, Chronic kidney disease, heart disease, Asthma/Chronic obstructive pulmonary disease, liver disease, cancer, pregnancy) or immunosuppressive therapy, that have been shown to increase the risk of acquiring SARS-CoV-2 were explored in the questionnaire.^2,8,13,15,19^ A prior SARS-CoV-2 diagnosis was self-reported by the participants and validated through the OHS personnel database in each hospital. The questionnaire was completed by each participant, with assistance offered to those participants requiring it.

### Data analysis

Data were exported from the REDCap^®^ online database for analysis using the IBM SPSS version 25.0 software (IBM SPSS, Chicago, Illinois) after cross-checking for completeness and accuracy. The means ± standard deviations were estimated for continuous data and counts and proportions were estimated for categorical data for the sociodemographic characteristics of the participants. The proportion of HCWs with either a SARS-CoV-2 PCR diagnosis or positive IgG antibodies, or both, were reckoned as cumulative incidence in the study. The cumulative incidence was disaggregated by sociodemographic and clinical factors.

The associations between the cumulative incidence and risk factors (sociodemographic and clinical) were explored using the Pearson χ^2^ test. We fitted both unadjusted and adjusted multi-variate logistic regression models to examine the independent risk factors for cumulative infection with SARS-CoV-2 among the HCWs in the study. Variable selection in the model analysis was guided by known risk factors reported previously in other studies.^8,13,15^ A p-value less than 0.05 was considered statistically significant.

### Ethical Considerations

The Walter Sisulu University Ethics Committee granted approval for the implementation of the study (Reference: 087/2020), as well as the Eastern Cape Provincial Department of Health and local hospitals ethics committee. Each participant provided written informed consent for the study. Participants’ rights to privacy and the confidentiality of clinical data were respected during and after the study. The research process followed the Helsinki Declaration and local institutional policy. All hard copies of materials used in the study were locked securely and soft copies were password-protected in the computer in the research office.

### Patient and public involvement

There was no public or patient involvement in the design, conduct or reporting of this research, as patients were not included. The healthcare worker participants were given their individual SARS-CoV-2 IgG results via cellular messaging. The main findings of the study will be shared with the respective hospital management teams.

## Results

A total of 1,309 HCWs participated in the study from both hospitals, 656 from Frere Hospital and 653 from Cecilia Makiwane Hospital. Eleven blood samples for SARS-CoV-2 IgG serology were missing or rejected by the laboratory and were excluded from the final analysis. Data for another three participants were excluded due to missing data on the main outcome measures. Data for 1,295 HCWs were included in the final analysis.

### Baseline characteristics of the participants (N = 1,295)

The participants were predominantly women (81.5%), black (78.7%), had undergone tertiary education (71.5%), and most had never smoked (91.0%). In terms of professional category, nurses predominated (44.8%), followed by support staff (28.8%) and medical doctors (13.6%). Most (77.1%) participants reported direct contact with patients with COVID-19 and had attended training on PPE use (79.4%) (Table 1).

**Table 1:**
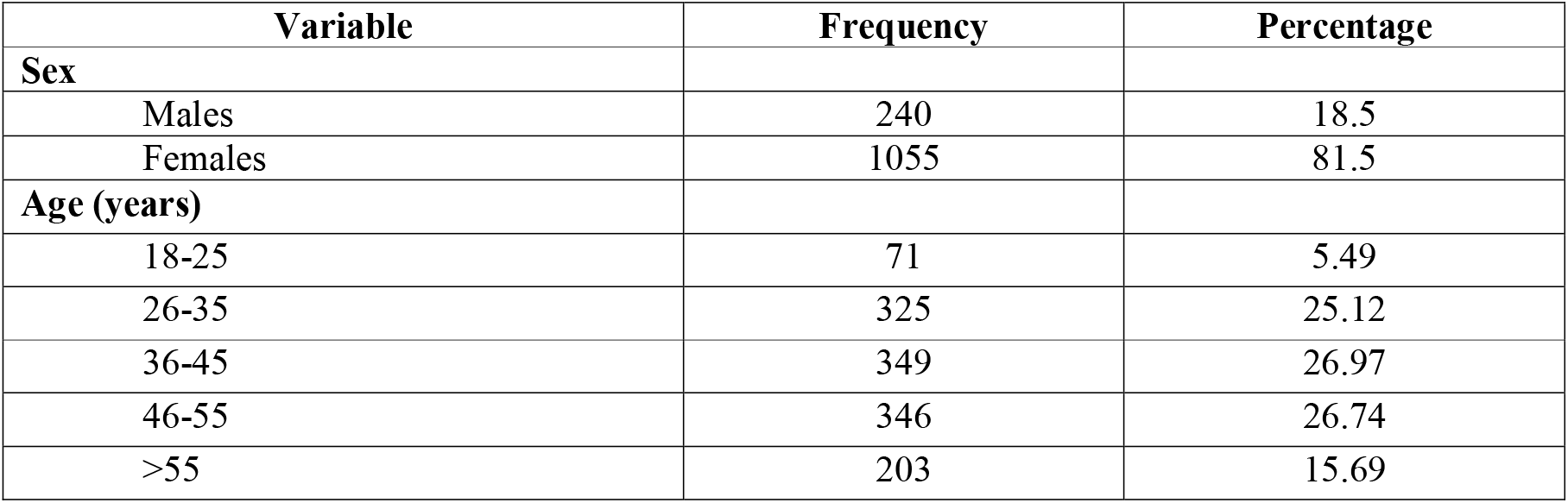
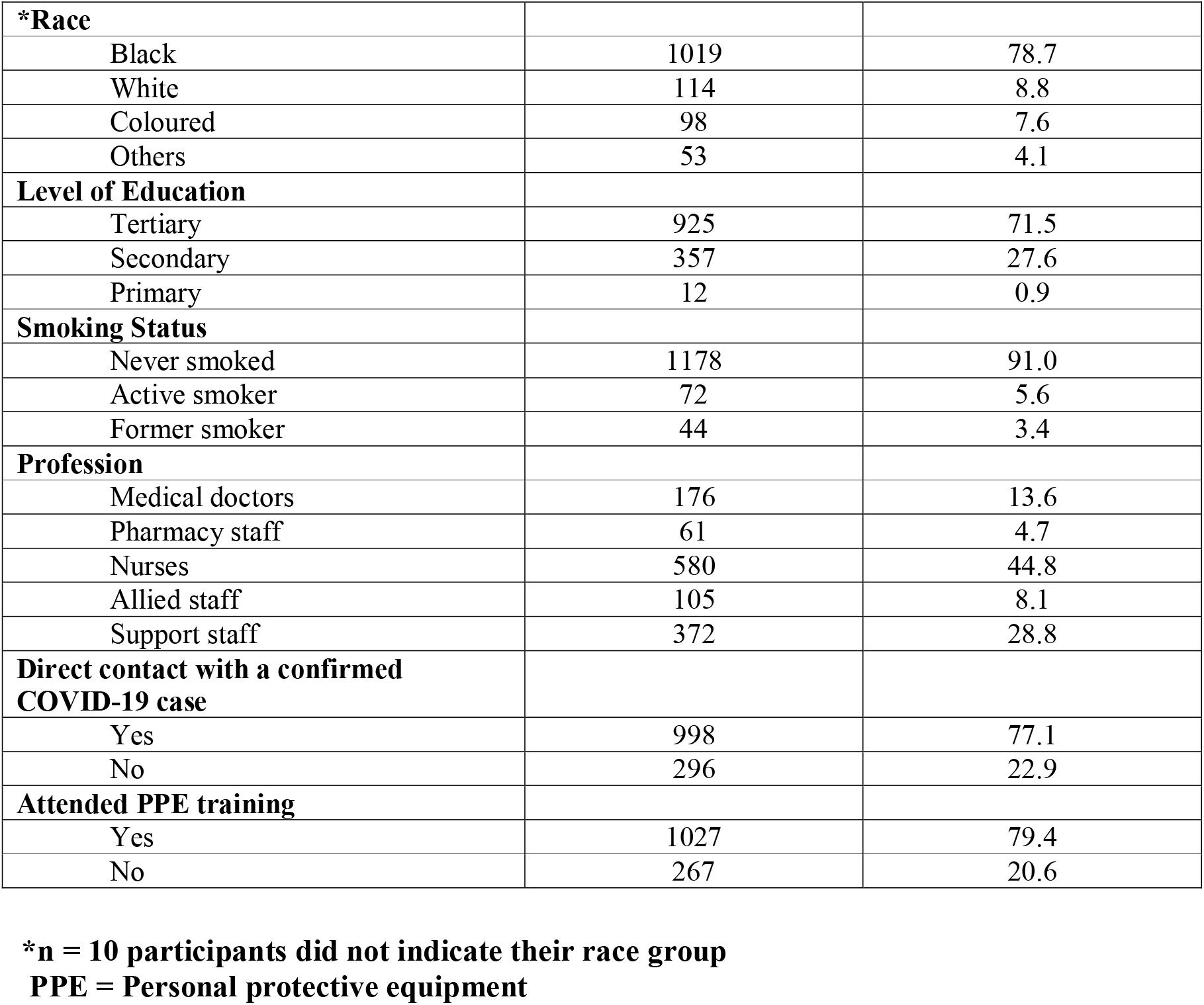
Baseline characteristics of the participants (n = 1,295)

| **Variable** | **Frequency** | **Percentage** |
| --- | --- | --- |
| **Sex** |  |  |
| Males | 240 | 18.5 |
| Females | 1055 | 81.5 |
| **Age (years)** |  |  |
| 18-25 | 71 | 5.49 |
| 26-35 | 325 | 25.12 |
| 36-45 | 349 | 26.97 |
| 46-55 | 346 | 26.74 |
| >55 | 203 | 15.69 |
| **PPE = Personal protective equipment** |  |  |
| **\*Race** |  |  |
| Black | 1019 | 78.7 |
| White | 114 | 8.8 |
| Coloured | 98 | 7.6 |
| Others | 53 | 4.1 |
| **Level of Education** |  |  |
| Tertiary | 925 | 71.5 |
| Secondary | 357 | 27.6 |
| Primary | 12 | 0.9 |
| **Smoking Status** |  |  |
| Never smoked | 1178 | 91.0 |
| Active smoker | 72 | 5.6 |
| Former smoker | 44 | 3.4 |
| **Profession** |  |  |
| Medical doctors | 176 | 13.6 |
| Pharmacy staff | 61 | 4.7 |
| Nurses | 580 | 44.8 |
| Allied staff | 105 | 8.1 |
| Support staff | 372 | 28.8 |
| **Direct contact with a confirmed COVID-19 case** |  |  |
| Yes | 998 | 77.1 |
| No | 296 | 22.9 |
| **Attended PPE training** |  |  |
| Yes | 1027 | 79.4 |
| No | 267 | 20.6 |

### SARS-CoV-2 cumulative incidence

SARS-CoV-2 infection was confirmed (PCR positive) in 390 participants (30.1%), and a SARS-CoV-2 IgG positive result occurred in 488 (37.7%), participants giving a cumulative incidence of 47.2% (611 HCWs). Of the 390 PCR positive cases, 123 (31.5%) were SARS-CoV-2 IgG negative at the time of study. One hundred and forty-six of 640 (22.8%) PCR negative cases were IgG positive indicating potentially false negative PCR tests or being tested at the incorrect time. The SARS-CoV-2 IgG picked up an additional 17.1% (n = 221) missed infections in this cohort (146 HCWs with negative PCR results and 75 who never tested) (Table 2).

**Table 2.** Confirmation of SARS-CoV-2 infection among the participants.

| **Variables** | **IgG Positive (n; %)** | **IgG Negative (n; %)** | **Total (n; %)** |
| --- | --- | --- | --- |
| PCR Positive | 267 (68.5) | 123 (31.5) | 390 (30.1) |
| PCR Negative | 146 (22.8) | 494 (77.2) | 640 (49.5) |
| Never tested | 75 (28.4) | 189 (71.6) | 264 (20.4) |
| Total | 488 (37.7) | 806 (62.3) | 1294 (100) |

IgG = Immunoglobulin G; PCR = Polymerase chain reaction; SARS-CoV-2 = Severe acute respiratory syndrome coronavirus-2

### Risk factors for SARS-CoV-2 infection among the HCWs

When examining sociodemographic and exposure risk factors for infection (Table 3), age, race, level of educational, smoking status, professional category, and work area were all significantly associated with SARS-CoV-2 infection (p < 0.05). Among the co-morbid conditions (Table 4), only Body Mass Index (BMI) was significantly associated with SARS-CoV-2 infection.

**Table 3.** Relationship between socio-demographic characteristics and SARS-CoV-2 by Pearson χ^2^ test.

| Variable | SARS-CoV-2 by PCR and/or IgG |  | p-values |
| --- | --- | --- | --- |
|  | Yes (%) | No (%) |  |
| **All** | n = 611 (47.2) | n = 683 (52.8) |  |
| **Sex** |  |  | **0.007** |
| Males | 95 (39.6) | 145 (60.4) |  |
| Females | 517 (49.0) | 538 (51.0) |  |
| **Age** |  |  | 0.628 |
| <45 years | 347 (46.6) | 397 (53.4) |  |
| >45 years | 346 (54.7) | 286 (45.3) |  |
| **Race** |  |  | **<0.001** |
| Black | 524 (51.4) | 495 (48.6) |  |
| White | 30 (26.3) | 84 (73.7) |  |
| Coloured | 29 (29.6) | 69 (70.4) |  |
| Others | 18 (34.0) | 35 (66.0) |  |
| **Level of Education** |  |  | **0.003** |
| Tertiary | 418 (45.2) | 507 (54.8) |  |
| Secondary | 191 (53.5) | 166 (46.5) |  |
| Primary | 02 (16.7) | 10 (83.3) |  |
| **Smoking Status** |  |  | **<0.001** |
| Never smoked | 580 (49.2) | 598 (50.8) |  |
| Active smoker | 17 (23.6) | 55 (76.4) |  |
| Former smoker | 14 (31.8) | 30 (68.2) |  |
| **Covid-19 exposure by Ward** |  |  | **0.008** |
| High risk | 151 (51.2) | 144 (48.8) |  |
| Medium risk | 265 (42.7) | 355 (57.3) |  |
| Low risk | 195 (51.5) | 184 (48.6) |  |
| **Profession** |  |  | **<0.001** |
| Medical doctors | 55 (31.2) | 121 (68.8) |  |
| Pharmacy staff | 28 (45.9) | 33 (54.1) |  |
| Nurses | 311 (53.6) | 269 (46.4) |  |
| Allied staff | 25 (23.8) | 80 (76.2) |  |
| Support staff | 192 (51.6) | 180 (48.4) |  |
| **Direct contact with a confirmed COVID-19 case** |  |  | 0.337 |
| Yes | 464 (46.5) | 534 (53.5) |  |
| No | 147 (49.7) | 149 (50.3) |  |
| **Attended PPE training** |  |  | 0.498 |
| Yes | 480 (46.7) | 547 (53.3) |  |
| No | 131 (49.1) | 136 (50.9) |  |

IgG = Immunoglobulin G; PCR = Polymerase chain reaction; PPE = Personal protective equipment; SARS-CoV-2 = Severe acute respiratory syndrome coronavirus-2.
Support staff = Administration/Management staff (51/98; 52.0%), General workers (31/61; 50.8%), Kitchen staff (23/33; 69.7%), Porters (06/15; 40.0%), Stores/Sales staff (0/5), Mortuary staff (4/5; 80.0%), Laundry staff (23/39; 59.0%). \*117 of the support staff did not indicate their duties.
Allied Workers = Radiology staff (9/37; 24.3%), Social workers (1), Physiotherapists (1), Dieticians (1), \*68 of allied workers did not indicate their duties.

**Table 4:** Relationship between co-morbidities and SARS-CoV-2 by Pearson χ^2^ test.

| Variables | Positive SARS-CoV-2 PCR and/ or IgG |  | p-values |
| --- | --- | --- | --- |
|  | Yes (%) | No (%) |  |
| **All** | n = 611 (47.2) | n = 683 (52.8) |  |
| **\*BMI** |  |  | **<0.001** |
| Underweight | 4 (57.1) | 3 (42.9) |  |
| Normal weight | 47 (26.7) | 129 (73.3) |  |
| Overweight | 121 (41.4) | 171 (58.6) |  |
| Obese | 434 (53.5) | 378 (46.6) |  |
| **Diabetes** |  |  | 0.076 |
| Yes | 56 (54.4) | 47 (45.6) |  |
| No | 555 (46.6) | 636 (53.4) |  |
| **Hypertension** |  |  | 0.246 |
| Yes | 119 (50.6) | 116 (49.4) |  |
| No | 492 (46.5) | 567 (53.5) |  |
| **HIV** |  |  | 0.300 |
| Yes | 40 (42.1) | 55 (57.9) |  |
| No | 571 (47.6) | 628 (52.4) |  |
| **TB** |  |  | 0.141 |
| Yes | 11 (34.4) | 21 (65.6) |  |
| No | 600 (47.5) | 662 (52.5) |  |
| **Chronic Kidney Disease** |  |  | 0.074 |
| Yes | 07 (29.2) | 17 (70.8) |  |
| No | 604 (47.6) | 666 (52.4) |  |
| **Heart Disease** |  |  | 0.496 |
| Yes | 15 (53.6) | 13 (46.4) |  |
| No | 596 (47.1) | 670 (52.9) |  |
| **Asthma/COPD** |  |  | 0.143 |
| Yes | 31 (39.2) | 48 (60.8) |  |
| No | 580 (47.7) | 635 (52.3) |  |
| **Liver Disease** |  |  | 0.169 |
| Yes | 06 (31.6) | 13 (68.4) |  |
| No | 605 (47.5) | 670 (52.6) |  |
| **Cancer** |  |  | 0.515 |
| Yes | 8 (40.0) | 12 (60.0) |  |
| No | 603 (47.3) | 671 (52.7) |  |

BMI = Body mass index; IgG = Immunoglobulin G; TB = Tuberculosis; SARS-CoV-2 = Severe acute respiratory syndrome coronavirus-2; COPD = Chronic obstructive pulmonary disease.

In an unadjusted logistic regression analysis (Table 5), female sex, coloured ethnicity, a primary education, active smokers, medical doctors and allied staff, use of public transport, and being overweight and obese were significantly associated with SARS-CoV-2 infection. However, in the adjusted logistic regression (Table 5), comorbidity with HIV, and being overweight and obesity were independently associated with SARS-CoV-2 infection. Individuals who were living with HIV were almost twice as likely to be infected with SARS-CoV-2 (Adjusted Odd Ratio [AOR] = 1.78; 95% Confidence Interval [CI]: 1.38-2.08). Individuals who were overweight were twice as likely to be infected with SARS-CoV-2 (AOR = 2.15; 95% CI 1.44-3.20). Similarly, those who were obese were slightly more likely to be infected with SARS-CoV-2 (AOR = 1.37; 95% CI 1.02-1.85).

**Table 5.** Adjusted and unadjusted logistic regression model showing risk factors for SARS-CoV-2 infection among HCWs.

| Variables | UOR (95%CI) | p-value | AOR (95%CI) | p-value |
| --- | --- | --- | --- | --- |
| **Sex** |  |  |  |  |
| Males | Ref |  | Ref |  |
| Females | 1.48 (1.11-1.79) | **0.007** | 1.09 (0.78-1.51) | 0.595 |
| **Race** |  |  |  |  |
| Others | Ref |  | Ref |  |
| Coloured | 0.48 (0.27-0.86) | **0.015** | 1.06 (0.55-2.06) | 0.848 |
| White | 1.44 (0.71-2.91) | 0.311 | 1.46 (0.69-3.07) | 0.313 |
| Black | 0.91 (0.91-1.81) | 0.789 | 1.28 (0.61-2.71) | 0.504 |
| **Level of Education** |  |  |  |  |
| Tertiary | Ref |  | Ref |  |
| Secondary | 4.12 (0.89-18.91) | 0.068 | 3.05 (0.62-14.85) | 0.166 |
| Primary | 0.71 (0.56-0.91) | **0.008** | 0.90 (0.67-1.22) | 0.509 |
| **Smoking Status** |  |  |  |  |
| Never smoked | Ref |  | Ref |  |
| Active smoker | 0.48 (0.25-0.91) | **0.026** | 0.65 (0.32-1.29) | 0.222 |
| Former smoker | 1.51 (0.65-3.48) | 0.334 | 1.77 (0.73-4.25) | 0.199 |
| **Profession** |  |  |  |  |
| Support staff | Ref |  | Ref |  |
| Allied staff | 2.34 (1.60-3.42) | **<0.001** | 1.92 (0.83-4.43) | 0.124 |
| Nurses | 1.25 (0.73-2.16) | 0.409 | 0.84 (0.35-1.99) | 0.693 |
| Pharmacy staff | 0.93 (0.71-1.19) | 0.545 | 0.88 (0.42-1.84) | 0.747 |
| Medical doctors | 3.41 (2.08-5.58) | **<0.001** | 1.52 (0.67-3.45) | 0.316 |
| **COVID-19 Exposure by Ward** |  |  |  |  |
| Low risk | Ref |  | Ref |  |
| Medium risk | 0.69 (0.53-0.90) | 0.006 | 1.19 (0.59-2.41) | 0.749 |
| High risk | 0.97 (0.71-1.32) | 0.883 | 0.88 (0.42-1.86) | 0.611 |
| **Direct contact with a confirmed COVID-19 case** |  |  |  |  |
| No | Ref |  | Ref |  |
| Yes | 0.88 (0.67-1.14) | 0.338 | 1.01 (0.75-1.36) | 0.928 |
| **Attended PPE training** |  |  |  |  |
| Yes | Ref |  | Ref |  |
| No | 1.09 (0.83-1.42) | 0.498 | 0.99 (0.74-1.33) | 0.996 |
| **Use of public transport** |  |  |  |  |
| No | Ref |  | Ref |  |
| Yes | 0.63 (0.51-0.79) | **<0.001** | 0.94 (0.69-1.17) | 0.444 |
| **BMI** |  |  |  |  |
| Underweight | - |  | - |  |
| Normal | Ref |  | Ref |  |
| Overweight | 3.15 (2.19-4.53) | **<0.001** | 2.15 (1.44-3.20) | **<0.001** |
| Obese | 1.62 (1.23-2.12) | **<0.001** | 1.37 (1.02-1.85) | **0.033** |
| **Diabetes** |  |  |  |  |
| No | Ref |  | Ref |  |
| Yes | 0.73 (0.48-1.09) | 0.131 | 0.85 (0.55-1.32) | 0.480 |
| **Hypertension** |  |  |  |  |
| No | Ref |  | Ref |  |
| Yes | 0.84 (0.63-1.12) | 0.246 | 1.08 (0.78-1.48) | 0.628 |
| **HIV** |  |  |  |  |
| No | Ref |  | Ref |  |
| Yes | 1.25 (0.81-1.19) | 0.301 | 1.78 (1.38-2.08) | **0.012** |

BMI = Body mass index; HCWs = Healthcare workers; PPE = Personal protective equipment; SARS-CoV-2 = Severe acute respiratory syndrome coronavirus-2; UOR = Unadjusted odds ratio; AOR = Adjusted odds ratio

## Discussion

This cross-sectional survey of 1 295 HCWs from two large referral hospitals in the Eastern Cape Province combined two diagnostic modalities (SARS-CoV-2 PCR and SARS-CoV-2 IgG antibodies) to estimate the cumulative incidence of SARS-CoV-2 infection. The study showed a high rate of SARS-CoV-2 infection (47.2%) after the first wave of COVID-19 among the HCWs in the region. This rate is double the official figures reported for doctors and nurses subsequent to the second wave in the Eastern Cape province (18.2-22.3%).^7^ The 30.1% SARS-CoV-2 PCR positivity is significantly higher than the pooled prevalence of 11% (95% CI; 7-15%) from a systematic review of 46 studies among HCWs worldwide.^20^ Similarly the 37.7% SARS-CoV-2 IgG seropositivity is higher than the pooled prevalence of 7% (95% CI; 4-11%) of 27,445 HCWs in the same review.^20^

In order to obtain reliable epidemiologic data on the infection rate with SARS-CoV-2 for strategic planning, a minimum of two or more data sources should be combined. Findings from this study demonstrate the importance of combining PCR results with antibody testing within a population to assess more accurately the cumulative incidence of SARS-CoV-2 infection. Neither of the modalities alone was accurate in estimating the infection rate in the study as reflected by the 31.5% of IgG negative results in HCWs who had been documented as SARS-CoV-2 PCR positive. These most likely represent cases of decay in the humoral immune response with IgG levels falling below the assay detection threshold over time. A study of the duration of SARS-CoV-2 IgG anti-nucleocapsid antibodies among 452 HCWs reported decline starting within 1 month after first positive PCR, with an estimated half-life of 85 days and 50% seronegative after 7 months.^21^ On the other hand, SARS-CoV-2 IgG testing identified 17.1% of participants with infections that had been missed by PCR. Two thirds (146/221) of these missed infections reported negative PCR tests. These likely represent false negative PCR results; suboptimal sample collections, or swabs that were taken before or after the peak of viral shedding.^22–24^ The other third (75/221) of the missed infections had never had a PCR test performed. These were likely asymptomatic infections or patients with mild symptoms that did not lead to PCR testing.

In terms of risk factors for SARS-CoV-2 infection among HCWs, the only significant risk factors in the adjusted multivariate logistic regression analysis were having an increased BMI (overweight or obese) and being HIV positive. While these factors have been reported as risks for infection among the general population in some reports^2,5,12,14,18^, this is the first time they have been linked in a specifically HCW population. Stratifying areas of work into low, medium and high risk for SARS-CoV-2 exposure did not identify significant differences in infection risk, contrary to findings by Iversen et al.^8^ There was also no difference in infection prevalence across different professions. These are important negative findings of this study, and contribute some insights into SARS-CoV-2 exposure and transmission in the hospital environment. Of interest for epidemiologic purposes are two pertinent questions. ‘Why did doctors and nurses working in designated COVID-19 clinical areas not experience higher infection rates than non-clinical staff?’ and ‘Did improved use of PPE in these designated clinical areas effectively level this risk?’

Despite a large proportion (80%) of HCWs having been trained on the use of PPE, and they confirmed that PPEs were available for use, there was no correlation with SARS-CoV-2 infection in the cohort. A prospective study of SARS-CoV-2 infections among 10,034 UK HCWs, showed a lower risk of infection among ICU clinical staff, suggesting that training on PPE and strict adherence to infection control protocols protected staff in high risk areas.^9^ While there were concerns about inadequate quantities and quality of PPE during the period prior to the study, there was never a total shortage of PPE for use in COVID-19 clinical areas in either of the two facilities. Another plausible explanation for the results could be the strict adherence to symptom screening of all staff in the COVID-19 clinical areas throughout the period. Prompt diagnosis and isolation of infected individuals will prevent further spread among HCWs in the same work areas^2,3^. Furthermore, it was not infrequent for COVID-19 cases to be diagnosed in the non-COVID-19 clinical areas, which could account for similarly high proportions of staff infection in low, medium and high-risk clinical areas.

Transmission of SARS-CoV-2 between HCWs in the common areas during tea and lunch breaks, when staff interact socially with or without masks was not measured in the study, but is quite probable to have occurred to some degree. It was hypothesised that taking shared or public transport to work would increase the risk of infection compared to solo vehicle transport, but this was not found to be significant. At the time of this study, there were no community seroprevalence data with which to compare our findings. During the second epidemiologic wave, Sykes et al. reported a seropositivity rate of 63% among blood donors from the Eastern Cape, the highest among four provinces sampled in the country in January 2021.^25^ This study only sampled 1,457 donors, a highly selected group of healthy volunteers from four provinces. It is therefore difficult to estimate the community prevalence at the time of our study. Notwithstanding, there is a strong possibility of a high-exposure environment outside of the hospitals in the region. A previous UK study found that having a household COVID-19 contact was the strongest risk factor for HCW infection [AOR 4.82; 95% CI 3.45–6.72].^9^

Being overweight or obese has been linked to increased susceptibility to SARS-CoV-2 infection, as well as to disease severity and increased mortality. A meta-analysis of 20 studies assessing obesity and risk of SARS-CoV-2 infection found an odds ratio of 1.46 (95% CI 1.30-1.65).^26^ Poorer outcomes for respiratory viruses in the obese had been described prior to SARS-CoV-2 with the H1N1 influenza pandemic.^27^ The mechanisms for the increased vulnerability to SARS-CoV-2 among the overweight and obese are complex. Obesity is associated with a pro-inflammatory phenotype and systemic low-grade inflammation.^27^ Obesity dampens and delays both the innate and the adaptive immune response to infection with reduced efficacy of B- and T-cell responses. Obesity is also associated with poorer response to vaccination, likely through the same immune dampening effects.^27^ This sample of HCWs revealed alarmingly high rates of being either overweight (22.7%) or obese (63.1%), which is a concern due to increased vulnerability to respiratory viral infections as well as the non-communicable disease risks linked such as type 2 diabetes mellitus, hypertension, cardiovascular diseases and certain cancers.^28^

There is epidemiological evidence for an increased susceptibility to SARS-CoV-2 with HIV infection. A systematic review and meta-analysis of almost 21 million people across multiple continents reported a risk ratio of 1.24 (95% CI 1.05-1.46) for SARS-CoV-2 infection among people living with HIV compared to those uninfected by HIV.^29^ The HIV prevalence of 7.3% in this cohort may be an underestimate, given the self-reported nature of the data and some infected individuals may not have been diagnosed. The estimated adult HIV prevalence in the local district is 13.6%, as a comparison.^30^ Data on CD4 cell counts and whether HIV infected HCWs were on antiretroviral therapy were not obtained in this study, but could have added more insights into the HIV-related risk. Like obesity, HIV is an important vulnerability to be managed among HCWs in relation to SARS-CoV-2 and other infections such as *Mycobacterium tuberculosis*.

## Strengths and limitations

This is the first reported study to have combined two diagnostic modalities to estimate the cumulative incidence of SARS-CoV-2 infection among HCWs in South Africa. Findings will inform IPC policies in the region. However, this study does have some limitations. Due to the pragmatic nature of the local policy relating to PCR testing for SARS-CoV-2, testing was largely limited to symptomatic staff, which would have missed some asymptomatic infections. HIV serology and CD4 counts were not tested, but relied on self-reporting of individual HIV status, which may likely underestimate the burden of HIV in the cohort.

## Conclusion

We report a high SARS-CoV-2 cumulative incidence of 47.2% after the first epidemiologic wave among HCWs from two referral hospitals in the Eastern Cape, South Africa. This is one of the highest reported in the literature and more than double that of the official figures for HCWs in the region. Being overweight or obese were significant risks for infection, and over 85% of HCWs fell into these categories. HIV infection was also associated with increased infection in the cohort. There were similar rates of infection across low, medium and high SARS-CoV-2 transmission risk areas, suggesting that significant transmission of infection occurred between colleagues or outside the workplace. Staff wellness programmes should address weight reduction and regular HIV testing and treatment, to mitigate vulnerabilities in this essential workforce.

## Data Availability

All data produced in the present study are available upon reasonable request to the authors

## Acknowledgements

The authors are grateful for the support and enthusiasm shown by the management and staff of the two hospitals toward the implementation of the study.

